## Supplementary figures and images for "Genetic requirement for *Esrp1/2* in vertebrate pituitary morphogenesis"

### Supplemental Figures

Figure S1

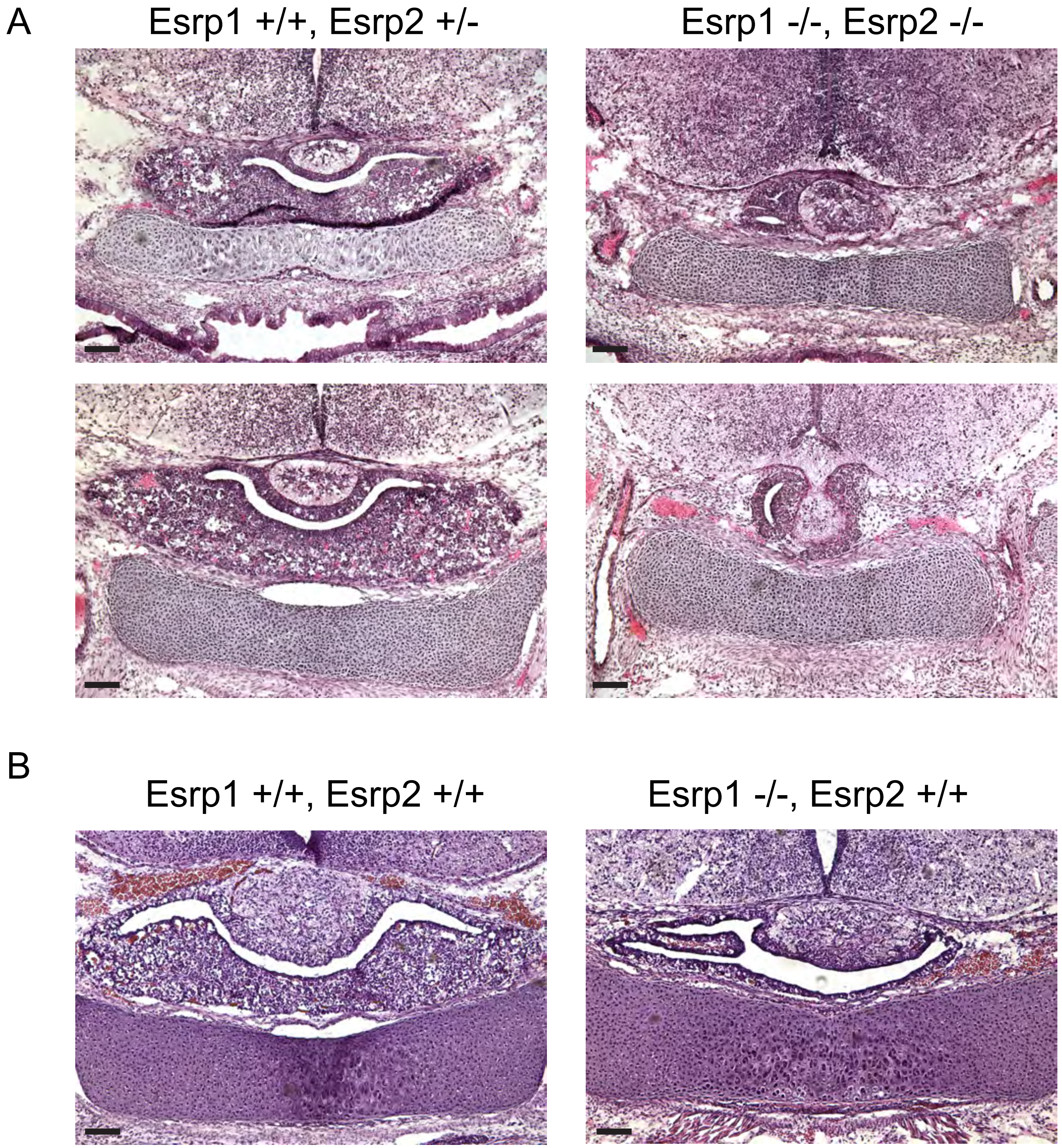

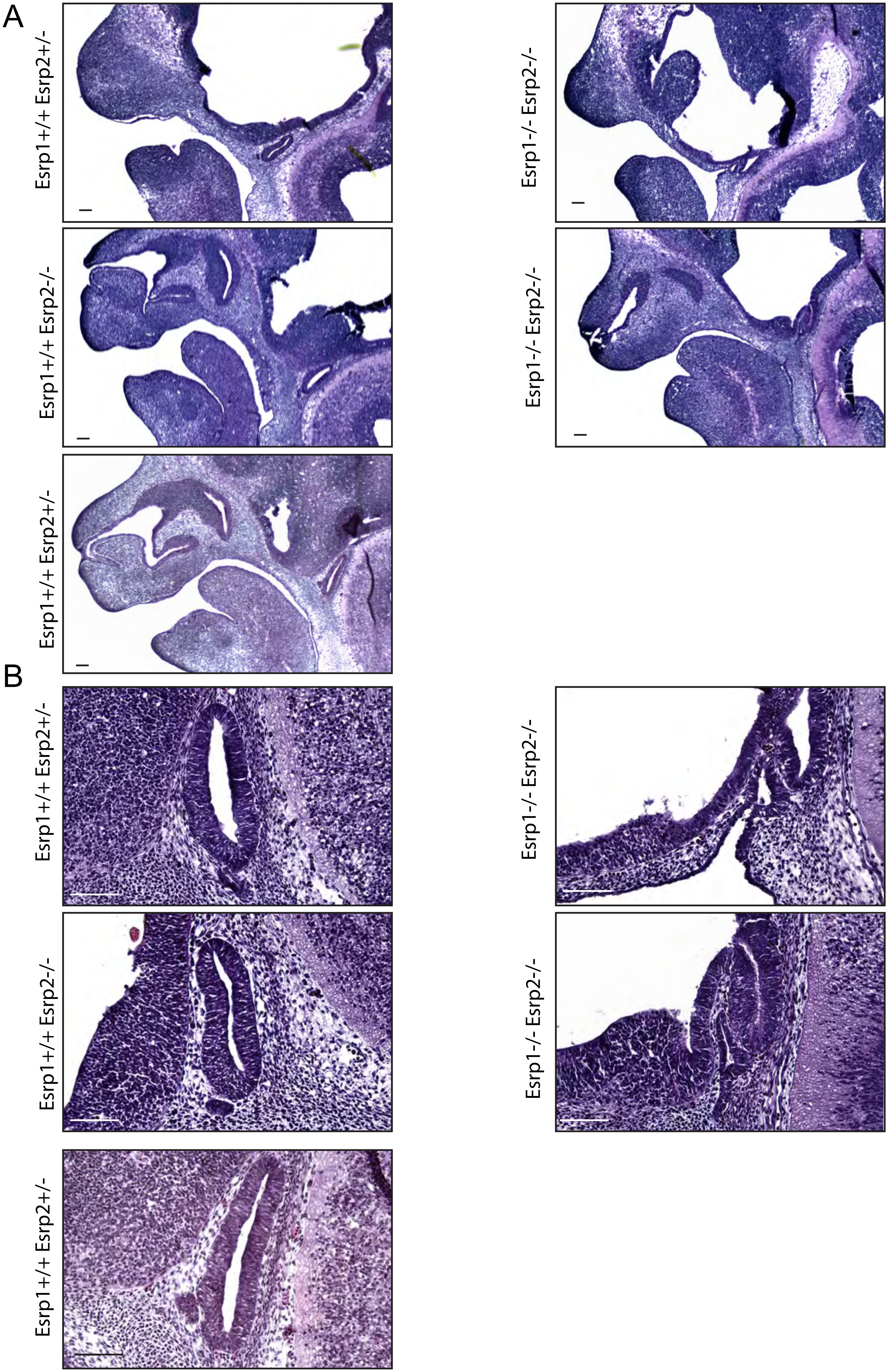
